# A profile analysis of peripherally inserted central catheters implanted over 10 years in a quaternary hospital

**DOI:** 10.64898/2026.04.22.26351492

**Authors:** Claudia Candido da Luz, Carolina Carvalho Jansen Sorbello, Estevão Araújo Epifânio, Caroline de Aguiar dos Santos, Simone Brandi, João Carlos de Campos Guerra, Nelson Wolosker

## Abstract

**Background:** Vascular access is essential in treating patients undergoing prolonged endovenous therapy such as chemotherapy, antibiotics, and parenteral nutrition. Since the 1990s, when PICCs (peripherally inserted central catheters) appeared, vascular access options have expanded significantly, revolutionizing the treatment landscape for all types of patients.

**Objective:** To analyze and describe the profile of the use of PICCs in a Brazilian quaternary hospital over 10 years with data collected by the infusion therapy team. Evaluating the number of PICCs implanted over the years, patients’ epidemiology and clinical characteristics, insertion details, associated complications, and the reason for removal.

**Methods:** A retrospective cohort study that employs a quantitative, non-experimental approach to classify and statistically analyze past events associated with 21,652 PICCs implanted from January 2012 to December 2021 in a quaternary hospital at São Paulo - Brazil. All the catheters were implanted, and the data was collected by a team of nurses specializing in infusion therapy. We analyzed the number of catheters implanted over the years, insertion characteristics, patients’ epidemiology and clinical data, possible associated complications, and the reason for removal. Statistical analyses were conducted using R software (version 4.4.1) and SPSS (version 29) for Windows (IBM Corp, Armonk, NY).

**Results:** During the specified period, 21,652 catheters were analyzed. The patients’ gender distribution was nearly balanced (48.2% versus 51.8%), and the average age was 66 years. Cardiovascular and metabolic issues were the most common comorbidities, and between 2020 and 2021, 29.3% of the sample tested positive for COVID-19. The most common location of hospitalization and implantation was the ward (31.6% - 44.2%), and the most used type of catheter was the Power Picc™ (83.9%). The estimated complication incidence density is 2.94 complications per 1,000 catheter-days. Almost all the PICCs (98,2%) were adequately located at the cavo-atrial junction after the first attempt, 82.2% of catheters were removed in the end of therapy, and the median duration of catheter use was 12 days.

**Conclusion:** PICCs are widely employed for drug infusion, with their use growing progressively due to specialized teams’ greater availability and training. The high efficiency of these devices with a relatively low risk of complications already observed in previous studies was reinforced by the findings of this study of more than 20,000 catheters.

## INTRODUCTION

Vascular access is essential in the treatment of patients undergoing prolonged endovenous therapy. Since the introduction of central venous catheters in the 1950s, the development of partially implantable catheters in the 70s, the advent of totally implantable catheters (TIC)(1) in the 80s, and finally, the use of PICCs (peripherally inserted central catheters) in the 80s/90s, the options for vascular access have significantly expanded, revolutionizing the treatment landscape for all type of patients(2,3).

PICCs are central catheters inserted through peripheral veins(4), commonly used for long-term endovenous treatments such as antibiotics administration, parenteral nutrition, and chemotherapy, among others(5). The insertion of PICCs is typically performed by specialized nursing teams or vascular surgeons in some medical settings. This procedure is generally performed without the need for an operating room or even on an ambulatory basis(6). It involves the use of local anesthesia, ultrasound-guided peripheral vein puncture(7), and confirmatory radiography to ensure proper placement before the catheter is approved for immediate use. The basilic, cephalic, and brachial veins are the most used for a puncture, allowing for safe central venous access for drug infusion through catheters exteriorized in the upper limbs(8).

Because this equipment is implanted in the arms, it provides several advantages over other central venous catheters(9). These benefits include reduced user exposure and embarrassment, increased comfort during treatment, and a lower risk of infection or catheter breakage. These advantages have led to the increasingly frequent use of such systems, especially for prolonged medication infusion and administering vesicant drugs. However, despite these many advantages, the catheter is not without complications(7,10), which occur infrequently. The most common issues include infections(11,12), Deep venous thrombosis (DVT) (13), and catheter malfunctioning.

In Brazil, the largest Latin American country with a population exceeding 200 million, the use of PICCs began in the 1990s(9) in neonatal ICUs. It quickly spread to other areas, including oncology (6,14) and pediatrics. Currently, the use of PICCs is widespread in both public and private healthcare systems, thanks to their high availability, the significant number of trained professionals, and the relative ease of implantation and management.(15)

Although PICC has become widespread over recent years, few studies with large casuistry have yet been done in Brazil, especially longitudinal studies that involve a substantial number of cases and long-term follow-up.

This study aimed to analyze the outcomes associated with the implantation of 21,652 PICCs between 2012 and 2021 in patients admitted to a quaternary hospital with a dedicated specialized nursing team. It analyzed the number of catheters implanted over the years, the demographic profile of the patients, details of hospital admissions, diagnoses of COVID-19 and other comorbidities, prophylaxis for DVT, catheter specifications and complications, and, finally, the length of time the device was used and the reasons for its removal.

## METHODS

### Design

This retrospective cohort study employs a quantitative, non-experimental approach to classify and statistically analyze past events from January 2012 to December 2021. The study received approval by the Medical Ethics Committee under CAAE number 65125022.6.0000.0071.

### Study site and data collection

The study was conducted at a quaternary hospital located in the southern part of São Paulo, Brazil. This private institution is recognized for its specialized services.

The Infusion Therapy Team (TTI) is responsible for inserting and monitoring PICCs and peripheral catheters inserted in patients with challenging venous access and Central Venous Catheters (CVC) placed by the medical team.

The TTI engages in numerous activities, including recording assessments, catheter insertions, and ongoing monitoring of these devices. A data form was completed for each assessment, whether it involves catheter insertion. This form captured information related to the patient, the catheter details about the insertion procedure, any complications encountered, and catheter removal. Notably, the form does not include any patient identification data.

After the team’s nurses had filled out the form, the information was entered into a database created using a Microsoft Office - Excel® tool. Two TTI monitors were responsible for managing and maintaining this database.

### Population/sample

Data relating to all consultations conducted by the TTI from January 2012 to December 2021 were analyzed. Additionally, we evaluated consultations requested for PICC® evaluation, even if the catheter was not inserted at the time. It resulted in a 21,652 sample.

### Procedures

The implantation procedures were performed in several care centers within a single hospital, including outpatient clinics, ward settings, surgical centers, and ICU beds. The PICCs used were mainly PowerPICC (BD, Becton, Dickinson, and Company - Switzerland), BioFLo (AngioDynamics - NY US), and Valved Groshong (BD, Becton, Dickinson, and Company - Switzerland), among others. All procedures were carried out by nursing staff specializing in infusion therapy.

The procedure consisted of peripheral venipuncture guided by portable ultrasound, particularly in the basilic, brachial, cephalic, and axillary veins. It was conducted using an aseptic technique and local anesthesia. The catheter was then advanced to the presumed central position (the atrio-caval junction), based on previously measured puncture points up to the height of T3/T5, where the atrio-caval junction is typically located. Finally, a flow and reflux test was performed, along with a confirmatory radiography, before the catheter was deemed ready for use.

The primary function of the PICC was to facilitate the infusion of medications such as antibiotics and chemotherapy. Once implanted and its position confirmed, the system was immediately ready for use.

The nursing team performed dressing changes during the patient’s hospital stay. They assessed the pouch, subcutaneous tunnel, and incisions for any signs of inflammation. The nursing team advised patients and their relatives to remain vigilant and keep changing the dressings at home. Patients and their companions were also informed about other possible warning signs, including fever, swelling pain, changes in coloration of the affected limb, and the importance of seeking medical assistance if these symptoms occurred.

At the end of treatment, the catheters were removed due to the inherent risks of complications, such as infections and DVT. Some catheters were removed earlier due to malfunctions or other complications.

### Follow-up

Patients were prospectively followed up until the removal of the catheter or until other outcomes occurred, including death.

We collected data on the number of catheters implanted during the period, the demographic profile of the patients, including age and biological sex, the length of time the device was used, the DVT prophylaxis used, the thrombosis risk and detection, and the diagnosis and place of hospitalization.

Additionally, we documented COVID test results and the presence of comorbidities, the type of catheter implanted, the number of insertion attempts, and other insertion characteristics. We also analyzed early and late complications, the reasons for withdrawal, and the final outcomes. The Padua-Caprini score categorized all patients to assess the risk of thrombosis, and a targeted Doppler ultrasound was performed on patients with suspected DVT.

### Data processing

Descriptive analysis utilized absolute frequencies and percentages for categorical variables, while means, standard deviations, medians, and quartiles were used for quantitative variables, along with the minimum and maximum values in the sample. To estimate incidence rates, Poisson regression models were used, with exposure time as offset. In the presence of missing data, the analysis was performed on the number of available cases. Statistical analyses were performed using R software (version 4.4.1) and SPSS (version 29) for Windows (IBM Corp, Armonk, NY).

## RESULTS

We evaluated 21,652 PICCs insertions on patients over 10 years, encompassing all age groups and various clinical conditions. Table 1 illustrates the distribution of procedures over the years.

**Table 1.**
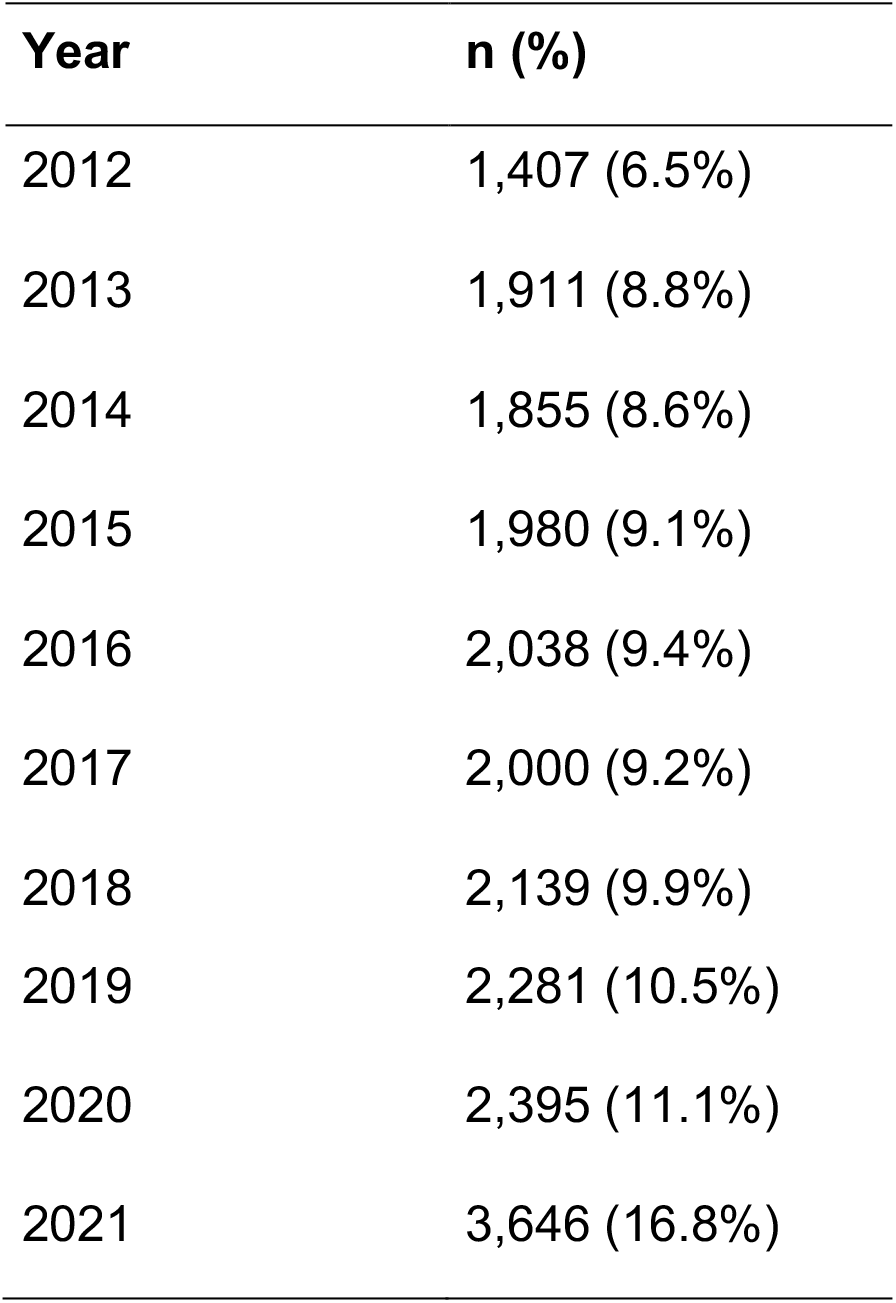
number of PICCs insertions over the years.

We observed a gradual increase in the number of PICCs implanted over the years. The patients’ characteristics indicated that the gender distribution was nearly balanced (48.2% versus 51.8%). The ages of the patients ranged from 0 to 109 years, with average age of 66 years and a median age of 73 years. We observed a predominance of older patients, with 75% of them being 53 years or older. The most common location for hospitalization was the ward (31.6%), followed by the Intensive Care Unit (ICU) (28%) and Semi-Intensive Care Unit (SICU) (27%). Specialized care areas (e.g., oncology, pediatrics, and intervention) cater to smaller patient populations. Regarding patient type, the majority were classified as clinical categories (76.4%). For further details, please refer to Table 2.

**Table 2.**
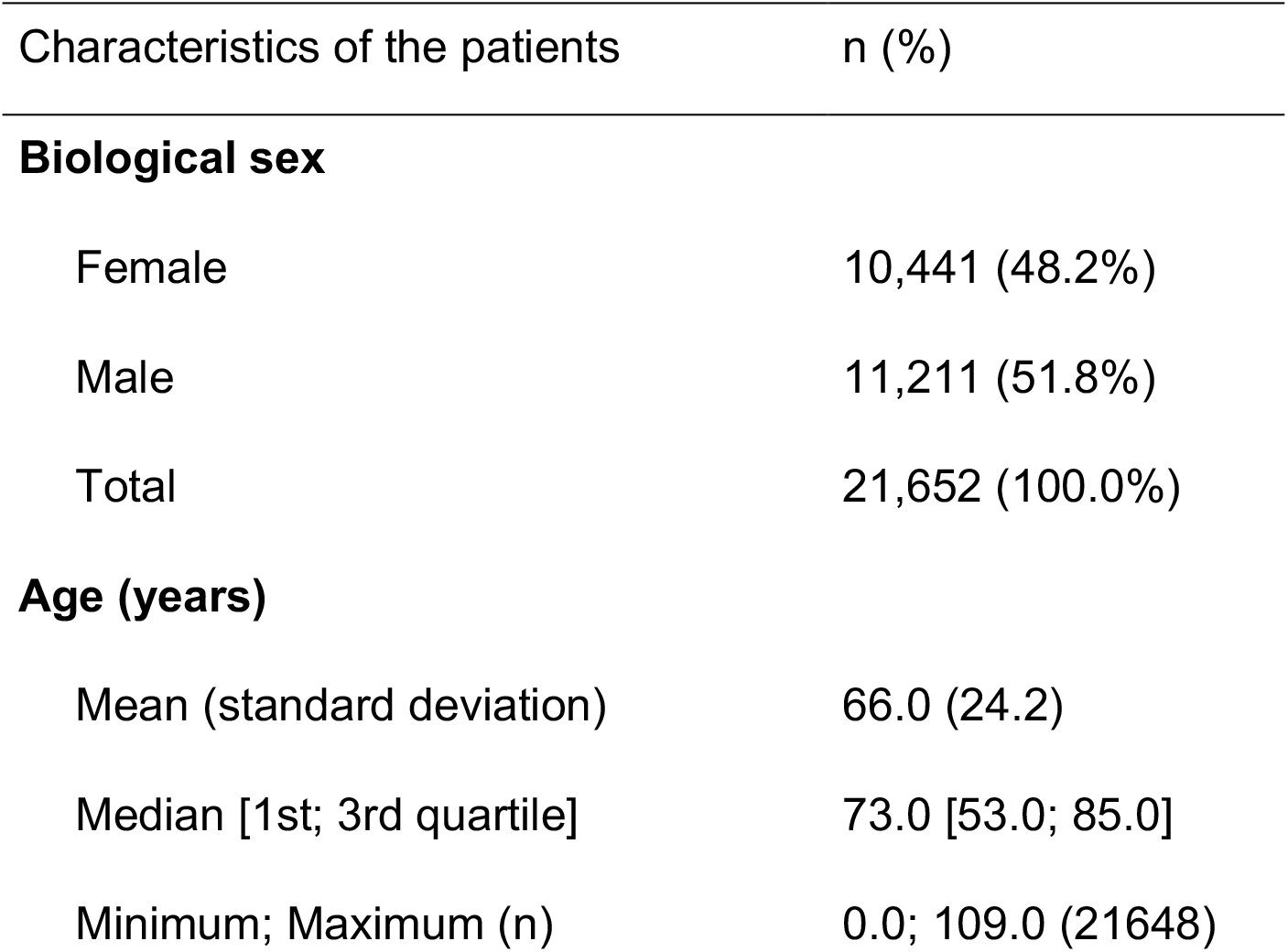
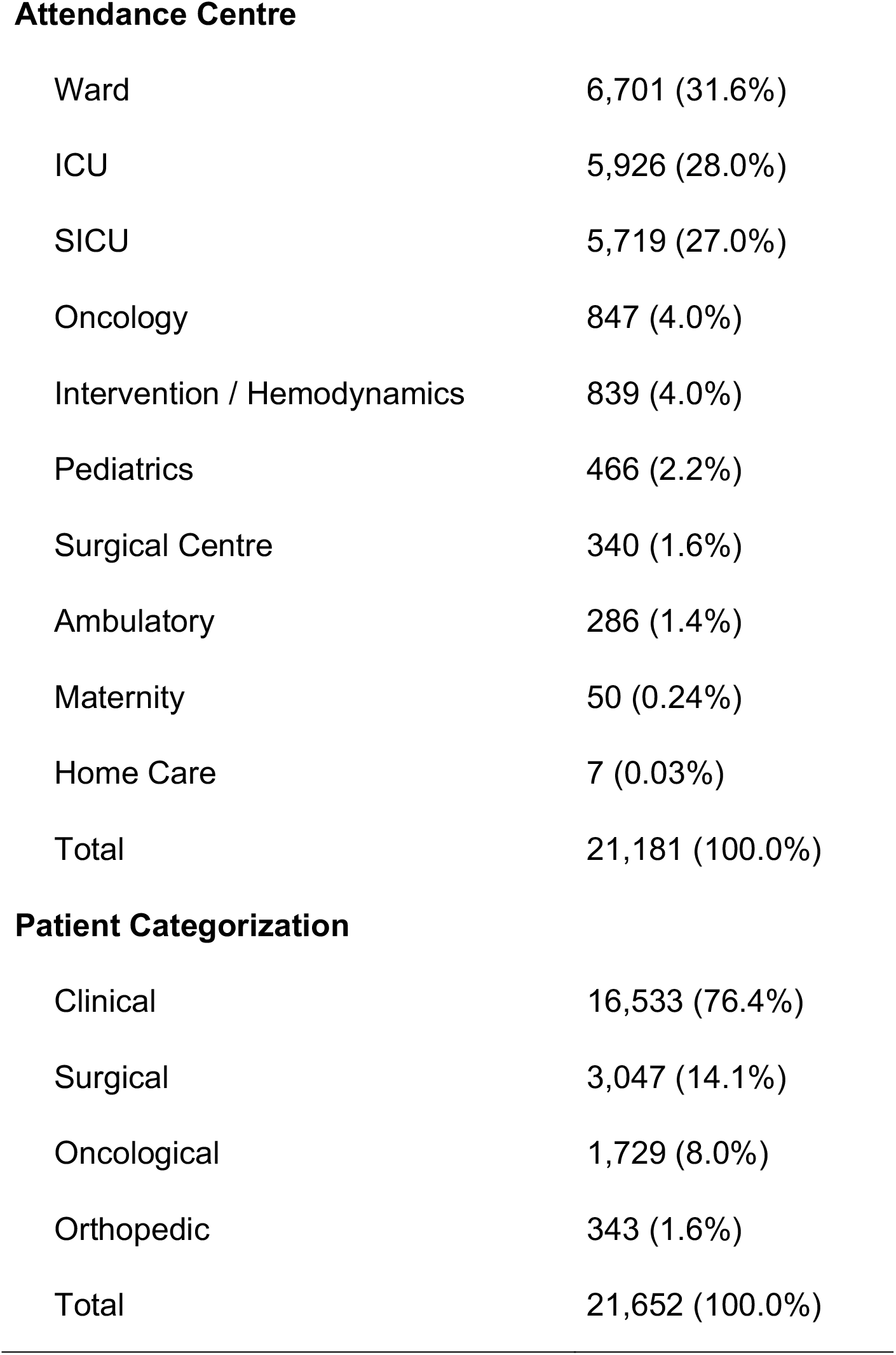
Characteristics of the patients studied.

| Characteristics of the patients | n (%) |
| --- | --- |
| **Biological sex** |  |
| Female | 10,441 (48.2%) |
| Male | 11,211 (51.8%) |
| Total | 21,652 (100.0%) |
| **Age (years)** |  |
| Mean (standard deviation) | 66.0 (24.2) |
| Median [1st; 3rd quartile] | 73.0 [53.0; 85.0] |
| Minimum; Maximum (n) | 0.0; 109.0 (21648) |
| **Attendance Centre** |  |
| Ward | 6,701 (31.6%) |
| ICU | 5,926 (28.0%) |
| SICU | 5,719 (27.0%) |
| Oncology | 847 (4.0%) |
| Intervention / Hemodynamics | 839 (4.0%) |
| Pediatrics | 466 (2.2%) |
| Surgical Centre | 340 (1.6%) |
| Ambulatory | 286 (1.4%) |
| Maternity | 50 (0.24%) |
| Home Care | 7 (0.03%) |
| Total | 21,181 (100.0%) |
| **Patient Categorization** |  |
| Clinical | 16,533 (76.4%) |
| Surgical | 3,047 (14.1%) |
| Oncological | 1,729 (8.0%) |
| Orthopedic | 343 (1.6%) |
| Total | 21,652 (100.0%) |

The comorbidities of 10,816 patients were assessed, revealing significant prevalence frequencies for cardiovascular and metabolic issues: 38.8% had hypertension, 22.2% were diabetic, and 16.2% suffered from dyslipidemia.

**Figure 1:**
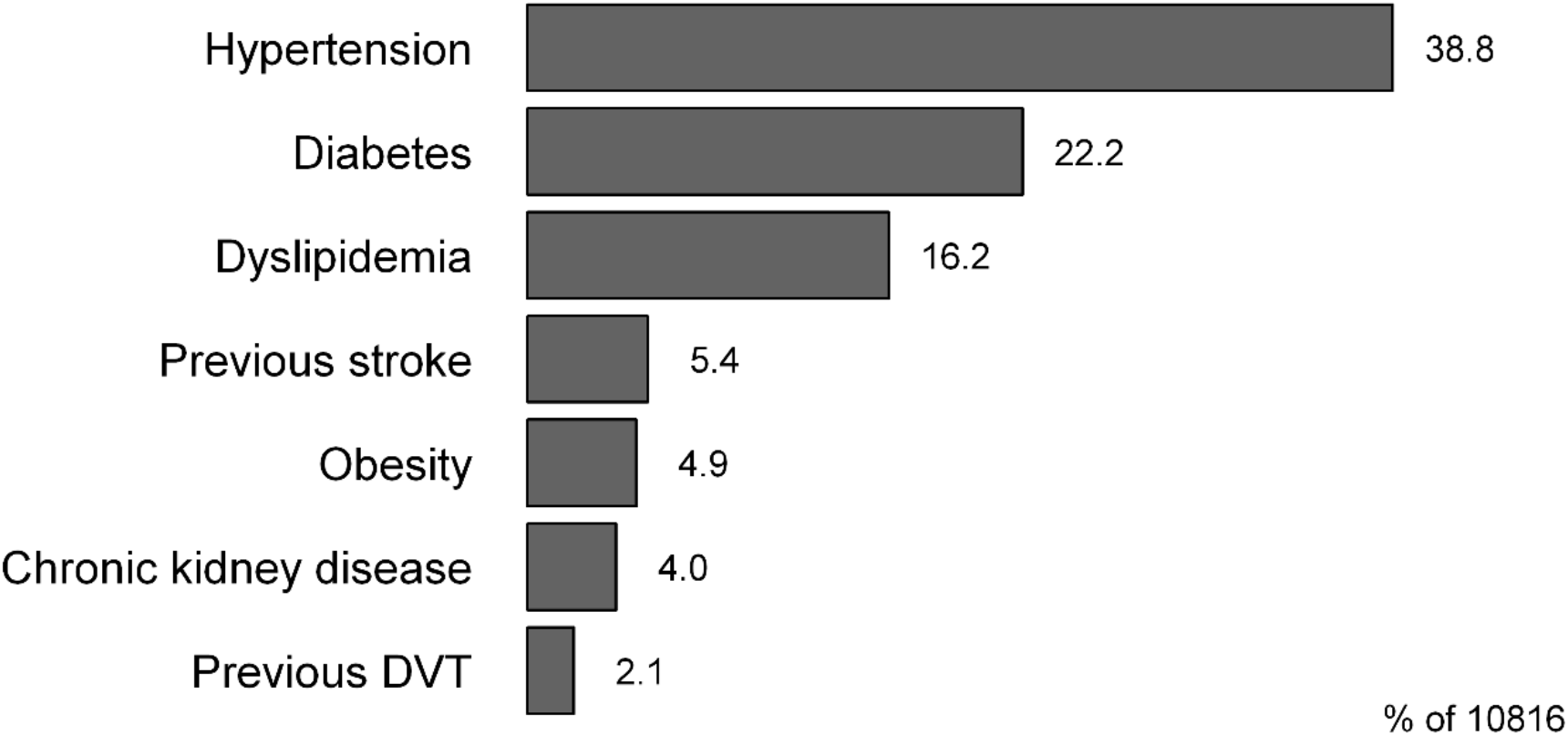
frequency of comorbidities.

During this period, another highly relevant health condition was COVID-19 infection. Testing for COVID-19 was conducted in 5,671 cases, with 1,659 tests returning positive, accounting for 29.3% of the total. The distribution of positive cases was as follows: 543 cases in 2020 (26.8%), and 1,116 cases in 2021 (30.6%).

Patients were also assessed for their risk, prevention, and diagnosis of DVT, as shown in Table 3. Most patients (92.6%) were considered at high risk of DVT, and pharmacological prophylaxis was the most common preventative measure used (63.7%). In fact, DVT was not detected in most cases, accounting for 98.6%.

**Table 3.**
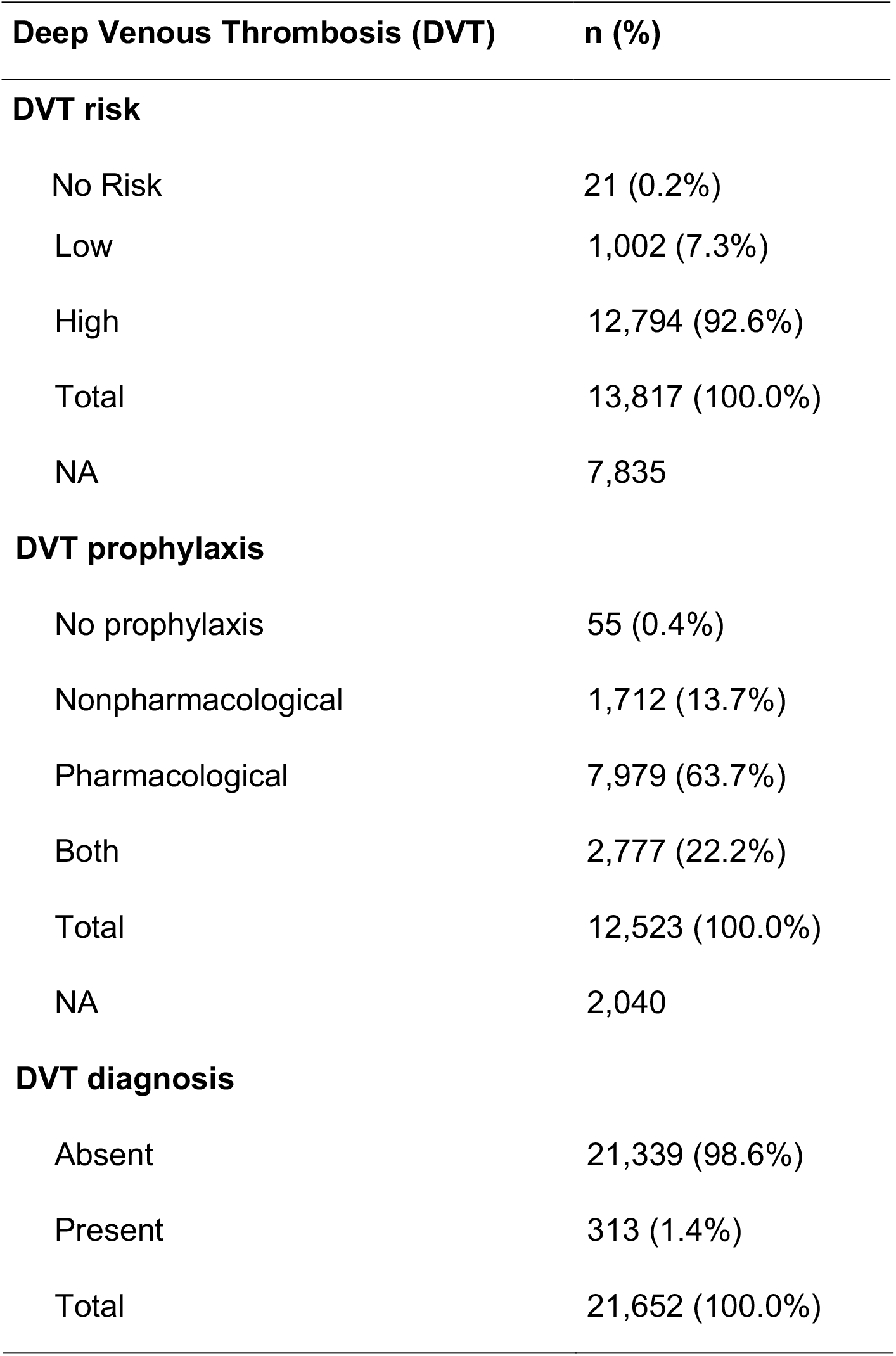
Risk, prophylaxis and diagnosis of deep venous thrombosis.

| Deep Venous Thrombosis (DVT) | n (%) |
| --- | --- |
| DVT risk |  |
| No Risk | 21 (0.2%) |
| Low | 1,002 (7.3%) |
| High | 12,794 (92.6%) |
| Total | 13,817 (100.0%) |
| NA | 7,835 |
| **DVT prophylaxis** |  |
| No prophylaxis | 55 (0.4%) |
| Nonpharmacological | 1,712 (13.7%) |
| Pharmacological | 7,979 (63.7%) |
| Both | 2,777 (22.2%) |
| Total | 12,523 (100.0%) |
| NA | 2,040 |
| **DVT diagnosis** |  |
| Absent | 21,339 (98.6%) |
| Present | 313 (1.4%) |
| Total | 21,652 (100.0%) |

Regarding the characteristics of PICC implantation, the most frequently used location for the procedure was the ward (44.2%). This was followed by the ICU (26.2%) and SICU (17.6%). The most used type of catheter was the Power Picc™ (83.9%), while other types were used in 15.2%. The final position of the catheter, as verified by the control X-ray, was adequate after the first attempt in 98.2% (cava-atrial junction). More detailed information is described in Table 4.

**Table 4.** Insertion and catheter characteristics.

| **Procedure Location** | **n (%)** |
| --- | --- |
| Ward | 6,236 (44.2%) |
| ICU | 3,700 (26.2%) |
| SICU | 2,487 (17.6%) |
| Surgical Center | 789 (5.6%) |
| Emergency Unit | 291 (2.1%) |
| Oncology | 249 (1.8%) |
| Ambulatory | 197 (1.4%) |
| Maternity | 122 (0.9%) |
| Intervention Center / Hemodynamic | 19 (0.1%) |
| Home Care | 5 (<0.1%) |
| Pediatrics | 3 (<0.1%) |
| Total | 14,098 (100.0%) |
| **Number of Lumens** |  |
| Mono | 1,645 (9.3%) |
| Double | 15,051 (85.2%) |
| Triple | 968 (5.5%) |
| Total | 17,664 (100.0%) |
| **Final Position (X-ray)** |  |
| Superior vena cava | 17,195 (98.2%) |
| Subclavian vein | 157 (0.9%) |
| Brachiocephalic vein | 110 (0.6%) |
| Atrium | 37 (0.2%) |
| Others | 15 (<0.1%) |
| Total | 17,514 (100.0%) |

Regarding complications, an absolute incidence of 4.9% was observed. The identified complications included events associated with malfunction or improper positioning of the device, as well as cases of breakage or loss, infection, and DVT, as shown in Table 5.

**Table 5.**
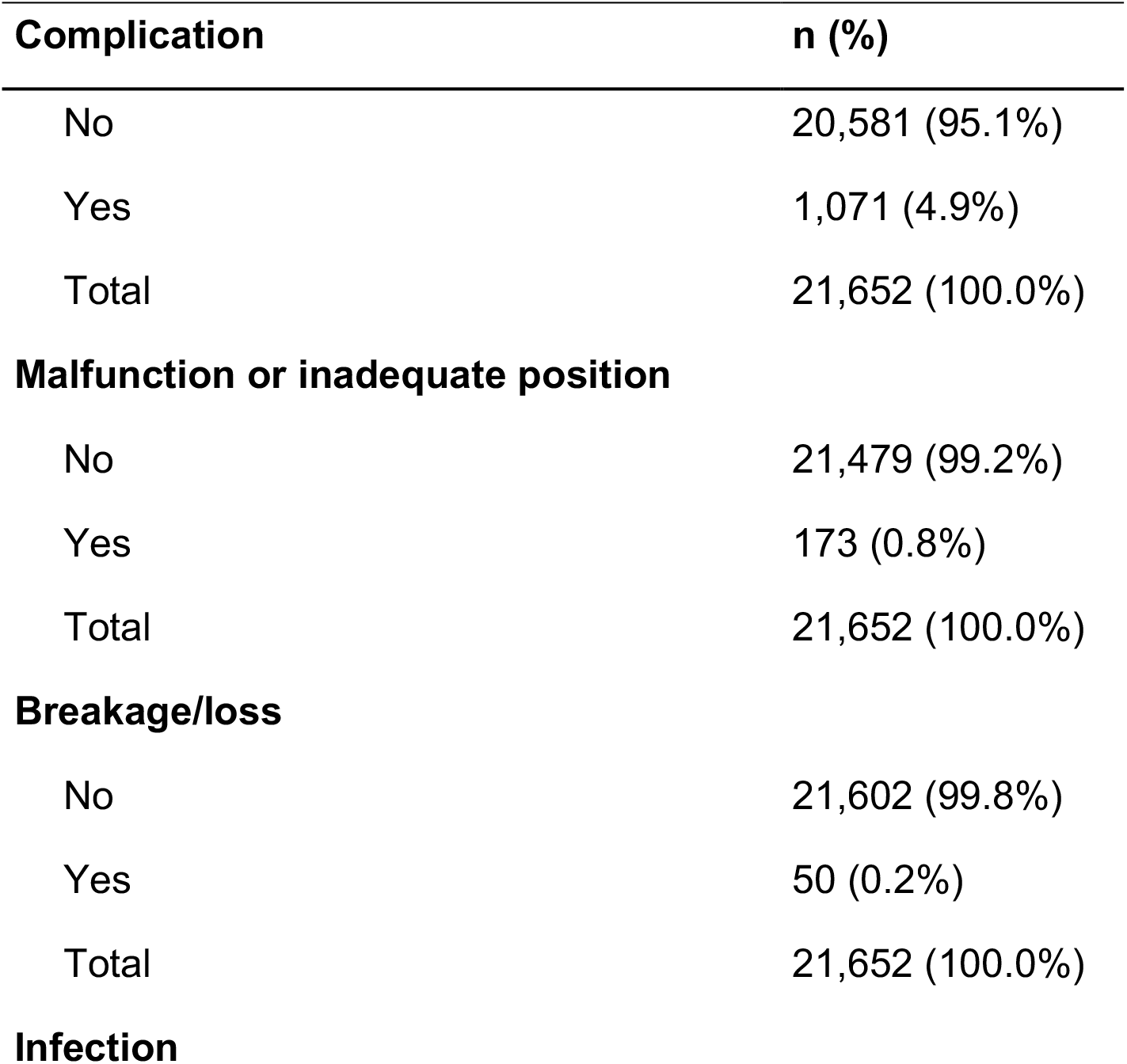
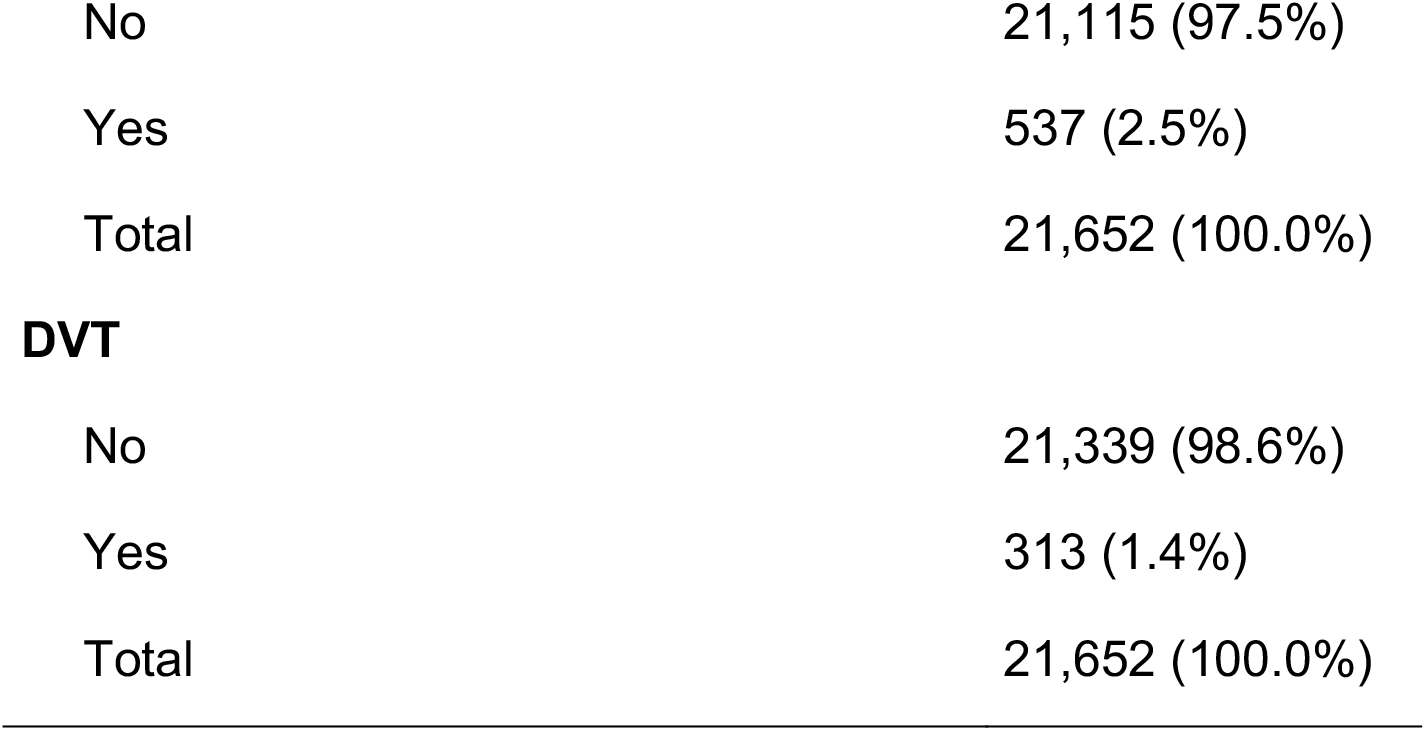
Complications.

| **Complication** | **n (%)** |
| --- | --- |
| No | 20,581 (95.1%) |
| Yes | 1,071 (4.9%) |
| Total | 21,652 (100.0%) |
| **Malfunction or inadequate position** |  |
| No | 21,479 (99.2%) |
| Yes | 173 (0.8%) |
| Total | 21,652 (100.0%) |
| **Breakage/loss** |  |
| No | 21,602 (99.8%) |
| Yes | 50 (0.2%) |
| Total | 21,652 (100.0%) |
| **Infection** |  |
| No | 21,115 (97.5%) |
| Yes | 537 (2.5%) |
| Total | 21,652 (100.0%) |
| **DVT** |  |
| No | 21,339 (98.6%) |
| Yes | 313 (1.4%) |
| Total | 21,652 (100.0%) |

The most frequent reason for catheter removal was the conclusion of therapy (82.2%) out of the 17,032 cases with this information available. Infection was cited as a reason for removal in 497 cases (2.9%). Additional reasons are shown in Table 6.

**Table 6.** Reasons for catheter removal.

| **Reason for Removal** | **n (%)** |
| --- | --- |
| End of therapy | 14,006 (82.2%) |
| Death | 1,887 (11.1%) |
| Infection | 497 (2.9%) |
| DVT | 277 (1.6%) |
| Malfunction or inadequate position | 173 (1.0%) |
| Inadvertent loss or removal | 50 (0.3%) |
| Catheter substitution | 38 (0.2%) |
| Others | 104 (0.6%) |
| Total | 17,032 (100.0%) |

We also analyzed the duration of catheter use, which ranged from one to 1,098 days, with a median of 12 days. Among the 12,363 cases where outcome information was available, 27.9% of patients were discharged with the PICC catheter. Regarding the discharge destinations, 66.9% were discharged to home care, 25.2% to ambulatory follow-up, 7.3% to another service, and 0.7% to other destinations.

## DISCUSSION

The implantation of a PICC for intravenous therapy has been a standard procedure in healthcare centers since the 1980s. This versatile type of catheter allows for the infusion of vesicant and irritating substances into the vascular system(5,15) for prolonged periods, helping to preserve patients’ peripheral venous systems. The procedure can be performed without complex infrastructure and carries low risks. While some studies have explored the use of PICCs in Brazilian hospitals, focusing on epidemiology, costs, and associated complications, no previous research has documented the experiences of a quaternary hospital with over 20,000 cases of PICCs implantation spanning 10 years.

The first mention of the PICC catheter dates back to the early 20th century in Germany when Dr. Forssmann(9) inserted a cannula into his own antecubital vein(16) and advanced it to the right atrium, confirming its position with radiographic imaging. This groundbreaking achievement earned him the Nobel Prize in Medicine in 1956. Starting in the 1950s, various types of catheters were developed, enabling treatments that would have otherwise been impossible(16). These include double-lumen catheters, semi-implantable catheters (such as Hickman and PermCath), fully implantable catheters, and eventually, the PICC catheter itself. Since then, the use of these devices has increased significantly, facilitating prolonged drug infusions and benefiting millions of patients worldwide(17).

In this study, we observed a consistent increase in the use of PICCs starting from 2012. This trend reflects a growing adoption and need for PICCs over the years. We identified three distinct phases in this trend:

- Phase One (2012 - 2015): This phase experienced an increase in PICC use, corresponding to the implementation period and increasing awareness of PICC usage. The number of PICCs rose from 1,407 in 2012 to 1,980 in 2015.
- Phase Two (2016 - 2018): In this period, the growth of PICC usage stabilized, with numbers gradually increasing from 2,038 in 2016 to 2,139 in 2018.
- Phase Three (2019 - 2021): This phase saw a sharp increase in the number of PICCs, reaching 3,646 in 2021 and accounting for 16.8% of the total. This surge coincided with the peak years of the COVID-19 pandemic in Brazil (2019 – 2021), when the demand for hospital admissions escalated dramatically across the country.

These findings highlight the changing role of PICCs in healthcare, particularly during critical periods such as the COVID-19 pandemic(18).

The use of PICCs has become a standard practice in our hospital. This shift can be attributed to increased awareness and training in PICC placement, advancements in technology and materials that have made these catheters more accessible and affordable, and improved catheter implantation techniques(19). Additionally, the growing need for long-term intravenous access due to various medical conditions and potential changes in healthcare policies or guidelines advocating for the use of PICCs have contributed to their increased usage over the years.

In almost half of the cases evaluated, catheters were implanted in a ward environment (44.2%). ICU and SICU were responsible for 43.8% of PICC insertions, showing the high frequency of catheter placement in critical care environments. Double-lumen catheters were the most common among the catheter types, reflecting a preference for balancing functionality and insertion complexity. The superior vena cava was identified as the most frequent final position for catheters (98.2%), demonstrating adherence to the guidelines established for the placement of central venous catheters. Furthermore, the main reason for catheter removal was the conclusion of therapy.

The typical patient profile recommended for prolonged drug therapy, especially chemotherapy(2), consists of older individuals with comorbidities. Our study confirms this finding, revealing a nearly balanced distribution of biological sex, with 11,211 males and 10,441 females. The average age of patients was 66 years, while the median age was 73 years, indicating a central tendency in the older adult population. This likely reflects a focus on age-related health conditions in the treatment procedures. Additionally, many patients presented with cardiovascular and metabolic risk profiles.

Most patients (76%) were classified in the clinical category, indicating a strong focus on non-surgical medical treatments. The majority of patients were located in the ICU, SICU, or general wards (primary care units). Surgical and oncological patients represented smaller subgroups within the total sample. Despite having fewer patients, oncology, pediatric, and surgical centers play a critical role in providing specialized care. These findings align with the catheter’s primary uses, which include administering antibiotics, parenteral nutrition, and chemotherapy.

The most part of our patients had hypertension (38.8%), diabetes (22.2%), and hypercholesterolemia (16.2%), which 49.1% of the sample that required a PICC presenting at least one of these three comorbidities. Patients with these conditions need preventive strategies centered on controlling hypertension, diabetes, and hypercholesterolemia to reduce cardiovascular risks. When patients have multiple comorbidities, they should be encouraged to seek multidisciplinary care, particularly those with chronic conditions like chronic kidney disease (CKD).

Since the end of 2019, Brazil has been registering cases of Covid-19(20), with a peak incidence occurring in 2021. This study shows that the rise in hospitalizations and number of patients with severe respiratory syndrome and its complications(21). In addition to maintaining urgent and emergency demands caused by other diseases,(22) has been accompanied by an increase in the need for central venous access. Interestingly, although there is no specific treatment for the virus, the use of catheters has proven essential for providing hemodynamic, nutritional, and antimicrobial support to critically ill patients.

The analysis of complications associated with the use of PICCs provided relevant data on the safety and efficacy of this device. In the period studied, the following were observed:

### PICC

The estimated complication incidence density is 2.94 complications per 1,000 catheter-days (95% CI: 2.76 to 3.12).

- Infection incidence density rate: 1.52 per 1,000 catheter-days (95% CI: 1.39-1.65).
- Thrombosis incidence density rate: 0.91 per 1,000 catheter-days (95% CI: 0.82-1.02).
- Malfunctioning incidence density rate: 0.51 per 1,000 catheter-days (95% CI: 0.44-0.59).

Compared to literature data:

### CVC

Complication incidence density ranges per 1,000 catheter-days(23).

- Infection incidence density rate: 4.8 per 1,000 catheter-days (95% CI: 3.4 - 6.6).
- Thrombosis incidence density rate: 2.7 per 1,000 catheter-days (95% CI: 1.0 - 6.2).
- Malfunctioning incidence density rate: 5.5 per 1,000 catheter-days (95% CI: 0.6 - 3.8).

### Port-a-Cath

Regarding port-a-cath catheters, an absolute incidence rate of 18.5% (95% CI: 16,7% - 20,5%) has been reported. (24).

- Infection incidence rate: 3.9 (95% CI: 3.1% - 5%).
- Thrombosis incidence rate: 3.2% (95% CI: 2.4% - 4.2%).
- Malfunctioning incidence rate: 7.9% (95% CI: 6.7% - 9.3%).

Infection was the main complication reported, occurring in 2.5% of patients and being responsible for 497 PICC removals. This rate is considered low for prolonged venous access devices, which indicates good insertion and maintenance practices, with good prevention protocols, such as regular dressing changes and the use of antiseptics.

The association between catheters for drug infusion and the occurrence of venous thrombosis(25) has been widely studied. In this article, we observed that drug prophylaxis with the correct indication (patients at high thrombotic risk according to the Caprini classification) could prevent DVT in 98.6% of the patients analyzed. This reinforces the importance of institutional protocols in implementing DVT prophylaxis.

The prevention protocols implemented in this study were associated with a low thrombosis rate, despite a largely high-risk population. The regular use of both pharmacological and non-pharmacological methods appears to have played a key role in achieving these outcomes. Nonetheless, further improvements could be made in standardizing risk assessment, ensuring long-term follow-up, and tailoring preventive strategies to individual patients. Enhancing these aspects may lead to even better patient safety and further reductions in thrombotic events.

Overall, PICCs are considered highly effective procedures with a low risk of complications, serving as an excellent alternative for safe infusion therapy, consistent with findings from previous studies worldwide. In addition, the use of PICCs has a positive economic impact by combining clinical efficiency with reduced costs compared to other venous access devices. The initial investment is offset by the lower incidence of complications, reduced length of stay and less need for reimplantation.

## LIMITATIONS

Due to the large number of patients studied over the years, with changes to the medical records system and the team dedicated to collecting data, some information was lost. However, this loss is minimal compared to the overall data.

As with any descriptive study, analytical inference takes a second place, as the primary objective is to describe the collected data.

Most catheters were implanted from 2019 onwards, so long-term follow-up for these patients is ongoing and will be reported in future studies.

## CONCLUSION

PICCs are widely employed for drug infusion in hospital centers, with their use growing progressively, especially following the COVID-19 pandemic. The majority of patients who require their implantation are elderly adults with multiple health conditions who are admitted for non-surgical treatment, especially in intensive care units.

Although this population is at a high risk of DVT, the incidence of thrombotic events is low. The primary reason for catheter removal is the completion of treatment, indicating that PICC lines are highly efficient and generally associated with a low risk of complications. Additionally, the rate of infection was also low, further supporting the safety profile of PICC use in this population.

## Data Availability

All data produced in the present study are available upon reasonable request to the authors

